# Factors influencing practice choices of early-career family physicians in Canada: A qualitative interview study

**DOI:** 10.1101/2023.05.07.23289626

**Authors:** Agnes Grudniewicz, Ellen Randall, M. Ruth Lavergne, Emily Gard Marshall, Lori Jones, David Rudoler, Kathleen Horrey, Maria Mathews, Madeleine McKay, Goldis Mitra, Ian Scott, David Snadden, Sabrina T. Wong, Laurie J. Goldsmith

## Abstract

**Purpose:** Comprehensiveness of primary care has been declining, and much of the blame has been placed on early-career family physicians and their practice choices. To better understand early-career family physicians’ practice choices in Canada, we sought to identify the factors that most influence their decisions about how to practice.

**Methods:** We conducted a qualitative study using framework analysis. Family physicians in their first 10 years of practice were recruited from three Canadian provinces: British Columbia, Ontario, and Nova Scotia. Interview data were coded inductively and then charted onto a matrix in which each participant’s data was summarized by code.

**Results:** Of the 63 participants that were interviewed, 24 worked solely in community-based practice, 7 worked solely in focused practice, and 32 worked in both settings. We identified four practice characteristics that were influenced (scope of practice, practice type and model, location of practice, and practice schedule and work volume) and three categories of influential factors (training, professional, and personal).

**Conclusions:** This study demonstrates the complex set of factors that influence practice choices by early-career physicians, some of which may be modifiable by policymakers (e.g., policies and regulations) while others are less so (e.g., family responsibilities). Participants described individual influences from family considerations to payment models to meeting community needs. These findings have implications for both educators and policymakers who seek to support and expand comprehensive care.

**Prior presentations:**

- “Factors influencing practice choices of early-career family physicians: A qualitative interview study.” North American Primary Care Research Group Conference (NAPCRG), Virtual, November 2021

Data also included in the following presentations on the broader study:

- “Addressing the need for greater primary care coverage: A discussion on primary care practice patterns”, Evidence Café, Ontario Ministry of Health & ICES. February 14, 2023.
- “The ‘kids’ are alright: Practice patterns among early-career family physicians and implication for primary care policy and workforce planning.” Family Medicine Grand Rounds, University of Ottawa. September 22, 2022.
- “Practice patterns among Early-Career Primary Care (ECPC) physicians and workforce planning implications: A mixed methods study. Summary of preliminary results.” British Columbia Ministry of Health and British Columbia General Practice Services Committee. February 25, 2022.

## Introduction

Despite the importance of family physicians, many people do not have access to one ^1, 2^ Concerns about declining access to comprehensive care ^3–7^ have generated speculation that the decline is a result of practice choices made by early-career physicians ^8–11^. Lack of access to family physicians providing comprehensive care has garnered attention from the media and the public, especially since the start of the COVID-19 pandemic ^12–14^. Family physicians have been pressured to see more patients and provide more services, leading many to retire early or step away from practice ^15^.

We know little about what influences early-career family physicians’ practice choices and what may be contributing to the move away from comprehensiveness. A recent study identified a decline in comprehensiveness across four Canadian provinces, but found that this decrease was not unique to new-to-practice physicians ^16^. As the next generation of providers, early-career family physicians’ practice choices will play an important role in the availability of comprehensive primary care going forward. An in-depth understanding of the reasoning behind their practice choices can inform policies and system reforms that would support a return to greater comprehensiveness in primary care delivery. In this paper, we answer the question, “what factors have influenced the practice choices of early-career family physicians in Canada?”

## Methods

We conducted a qualitative interview study using framework analysis ^17–19^, to identify factors influencing practice choices of early-career physicians. This study is part of a larger, mixed-methods project examining the practice patterns of early-career family physicians and residents in Canada ^20^. We received research ethics board approval from the Simon Fraser University Office of Research Ethics with harmonized approval from partnering universities.

## Data Collection

We recruited participants in three Canadian provinces – British Columbia, Ontario, and Nova Scotia – who were in their first 10 years of family practice. A description of the Canadian context is provided in Box 1. Chosen provinces vary with respect to geographic context, population characteristics, and payment and practice models. Invitations to participate were sent through family medicine residency program alumni email lists, and via Twitter, Facebook, and posters at research conferences.

Individuals interested in the study completed an online screening survey of demographic and practice-related characteristics. Maximum variation sampling of survey respondents enabled diverse participation across provinces, gender, years since residency, relationship status, dependents, practice setting, and specialized training. We aimed for approximately 20 participants from each province to ensure a sample size that allowed for wide-ranging practice experiences.

#### Box 1. Canadian Family Medicine Context

● Funding and delivery of primary care falls under provincial/territorial jurisdiction.
● Primary care models vary across the country and within provinces and territories.
● Most family physicians are private providers and are paid fee-for service.
● Capitation, blended, and salary payment models also exist and vary by province.
● To become a family physician, medical school graduates need to complete a two-year family medicine residency.
● Family physicians play the role of gatekeeper to specialist physicians in the health care system.

Data were collected through semi-structured interviews addressing topics about current practice characteristics, personal and professional influences, training experiences, the policy environment, and considerations for possible future practice changes (see Appendix for interview guide). Interviews were conducted by phone or videoconference by a single female researcher in each province, audio recorded, and lasted 45-60 minutes. Interviewers (ER, LJ, MMcK) had graduate-level training in qualitative research and were supervised by PhD researchers (AG, EM, LJG) with extensive experience in qualitative health services and primary care research. Interviewees received an honorarium to compensate for their time. Interview recordings were professionally transcribed.

## Data Analysis

Our analytic framework consisted of a subset of codes from the larger project codebook relevant to our research questions (see Box 2 for codebook development). The codes included current practice characteristics, and influential training, professional, and personal factors. All interview transcripts were coded using this analytic framework (i.e., the “indexing” step of framework analysis). Each transcript was then summarized for each component of the analytic framework (i.e., the “charting” step of the framework analysis). To ensure a consistent approach to summarizing and populating individual cells in the framework matrix, the qualitative lead (AG) and three members of the research team developed an analytic dictionary.

#### Box 2. Codebook Development Process

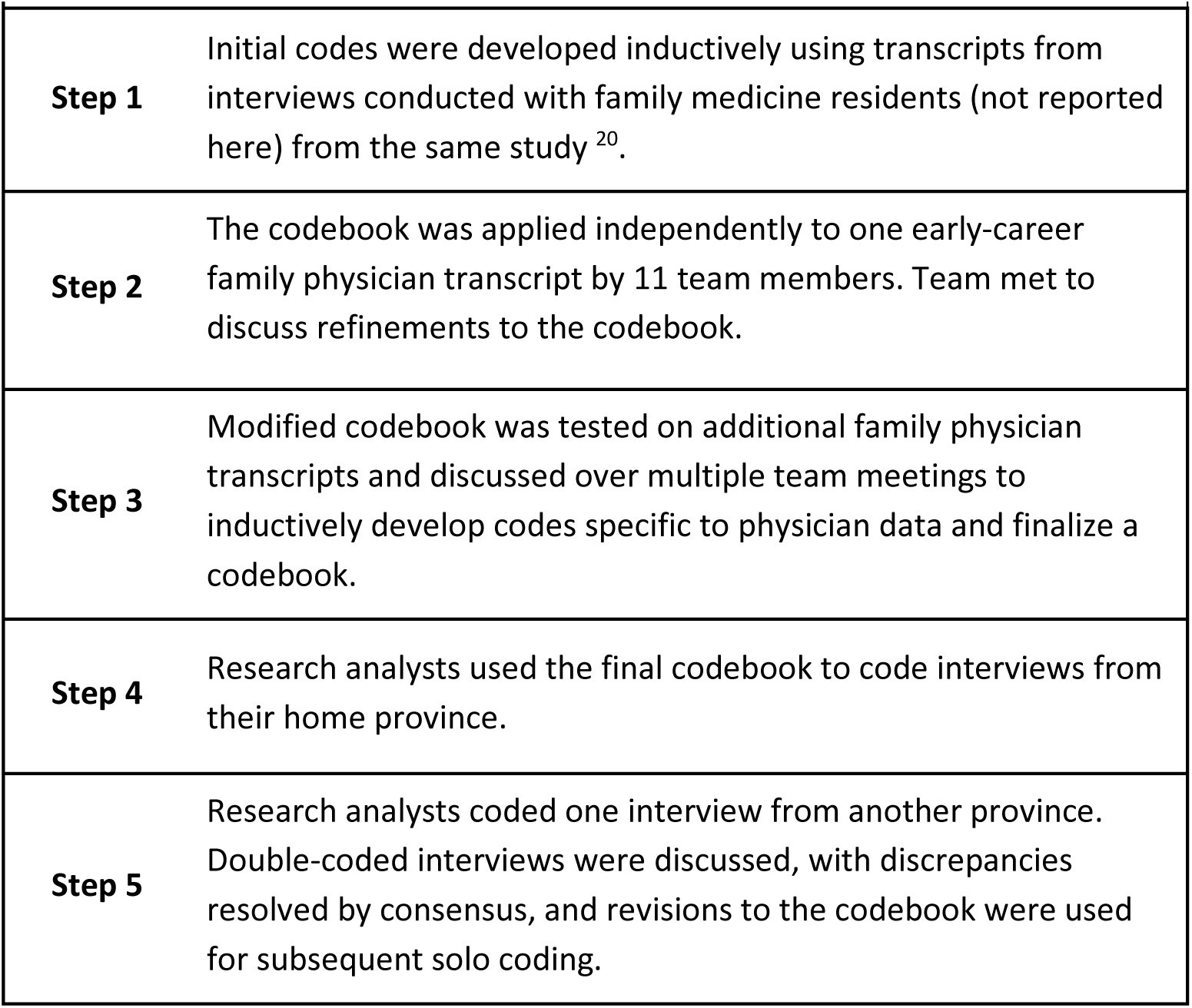

The analysis resulted in four matrices containing cross-sectional summaries for all study participants. The first matrix was concerned with four key practice characteristics being influenced (scope of practice, practice type and model, location of practice, and practice schedule and work volume). The remaining matrices addressed a specific category of influences (training, professional, personal). We then created interim matrices and tables that enabled us to explore the relationships between the characteristics and influences (i.e., final step of “mapping and interpretation”). We identified themes that summarized how each practice characteristic was influenced, together with illustrative quotes.

## Results

We present participant and practice characteristics, followed by the factors that influence practice choices. We use the term ‘community-based practice’ to refer to primary care provided to a general population in a clinic setting. We use ‘focused practice’ to refer to one or more specific clinical areas that make up a part, or all, of a family physician’s practice outside a traditional community-based practice ^21^.

### Participant Characteristics

We interviewed 63 early-career family physicians between April and October 2019. Our sample contained slightly more women than men, with around half having dependent children, and most having trained in Canada (Table 1).

**Table 1.** Demographic characteristics for the full study sample (n=63)

| **Province** | **n (%)** |
| --- | --- |
| British Columbia | 23 (36.5) |
| Ontario | 18 (28.6) |
| Nova Scotia | 22 (34.9) |
| **Gender\*** | **n (%)** |
| Woman | 36 (57.1) |
| Man | 26 (41.3) |
| Prefer not to answer | 1 (1.6) |
| **Relationship status** | **n (%)** |
| Single/Divorced/Separated/Widowed | 22 (34.9) |
| Married/Common-law/Life Partner | 39 (61.9) |
| Other | 1 (1.6) |
| Prefer not to answer | 1 (1.6) |
| **Dependents** | **n (%)** |
| Child(ren) | 34 (54.0) |
| Adult(s) | 2 (3.2) |
| Both | 1 (1.6) |
| None | 26 (41.3) |

| **Location of medical school** | **n (%)** |
| --- | --- |
| Canada | 48 (76.2) |
| Outside of Canada | 15 (23.8) |
| **Years since graduation from residency** | **n (%)** |
| 1-3 | 31 (49.2) |
| 4-6 | 20 (31.7) |
| 7-9 | 12 (19.0) |

\* Participants were given the options of “non-binary/third gender,” and “prefer not to self-describe” but these were not selected.

### Practice Characteristics

Participants worked in both community-based practices and focused practices, with almost a third working in locums (Table 2).

**Table 2.** Practice characteristics for the full study sample (n=63).

| **Practice type** | **n (%)** |
| --- | --- |
| Solely community-based practice | 24 (38.1) |
| Solely focused practice | 7 (11.1) |
| Both community-based and focused practices | 32 (50.8) |
| **Locuming** | **n (%)** |
| Locuming in community-based or focused practices | 20 (31.7) |
| Not locuming | 38 (60.3) |
| Unknown/Unclear* | 5 (7.9) |
| **Days per week in community-based practice** | **n (%)** |
| 0 days per week | 7 (11.1) |
| ≤ 2 days per week | 10 (15.9) |
| 2 to 4 days per week | 21 (33.3) |
| ≥ 4 days per week | 22 (34.9) |
| Unknown/Unclear* | 3 (4.8) |

\* Was not described during the interview by the participant.

Although our interview guide did not include structured questions about practice characteristics, 49 of the 56 physicians who worked in community-based practice described their practice characteristics (Table 3). Most participants worked in a group practice, at one site, and with other health professionals.

**Table 3.** Practice characteristics described by early-career family physicians in community-based practices (n=49).

| **Practice model** | **n (%)** |
| --- | --- |
| Solo practice | 2 (4.1) |
| Group practice | 41 (83.7) |
| Both solo and group practices | 3 (6.1) |
| Unknown/Unclear* | 3 (6.1) |
| **Number of practice sites** | **n (%)** |
| Single family practice site | 35 (71.4) |
| Multiple family practice sites | 14 (28.6) |
| **Working with other health professionals** | **n (%)** |
| Yes | 47 (95.9) |
| No | 1 (2.0) |
| Unknown/Unclear* | 1 (2.0) |
| **Working with other health professionals, by type of professional†** | **n (%)** |
| Family physician | 42 (85.7) |
| Specialist physician | 12 (24.5) |
| Nurse | 31 (63.3) |
| Pharmacist | 5 (10.2) |
| Other | 20 (40.8) |
| Unknown/Unclear* | 7 (14.3) |
| **Payment<sup>††</sup>** | **n (%)** |
| Fee for Service (FFS) and Enhanced FFS | 22 (44.9) |
| Salary | 21 (42.8) |
| Blended Capitation | 4 (8.2) |
| Supplements | 4 (8.2) |
| Incentives | 2 (4.1) |
| Other (e.g., hourly/daily/sessional rates) | 7 (14.2) |
| Unknown/Unclear* | 4 (8.2) |

\* Was not described during the interview by the participant. † An individual physician may work with more than one type of other health professional. †† An individual physician may be compensated by more than one payment model.

### Factors Influencing Practice Choices

Our analysis identified four key practice characteristics: 1) practice type and model, 2) scope of practice, 3) location of practice, and 4) practice schedule and work volume. These were influenced by three categories of influential factors: training, professional, and personal (see Table 4 for a visual overview of the data).

**Table 4.** Practice characteristics and the factors that influence them.

|  | Training influences |  |  | Professional influences |  |  |  |  |  |  |  |  |  | Personal influences |  |  |  |  |  |
| --- | --- | --- | --- | --- | --- | --- | --- | --- | --- | --- | --- | --- | --- | --- | --- | --- | --- | --- | --- |
|  | Medical school | Residency | Additional training | Early career experiences | Acquisition & maintenance of skills | Policies & regulations | Practice management responsibilities | Desire for control | Meaningful & valued work | Confidence with service provision | Challenges in rural settings | Offer better access to care | Remuneration | Financial considerations & pressures | Family & relationships | Gender | Personality & personal interests | Work-life balance | Community |
| Practice type & model | • | • |  | • | • | • | • | • | • |  |  |  |  | • | • | • | • | • |  |
| Scope of practice |  | • | • | • |  |  |  |  | • | • |  |  |  |  | • | • | • |  |  |
| Location of practice | • | • |  |  |  | • |  |  | • |  | • |  |  |  | • |  | • |  | • |
| Practice schedule & work volume |  |  |  |  | • |  |  |  |  |  |  | • | • | • | • |  |  | • |  |

\* A dot represents the presence of a relationship between a practice characteristic and influential factor as described by our participants.

**Table 5.** Participant perspectives on influences on practice type and model.

| Influence | Quotes |
| --- | --- |
| **Training influences on choice of practice type or model** |  |
| Medical school | "And I got to see how all these different GPs could find their own niche. Some of them liked to do more mental health, and some of them did endoscopy or GP anesthesia. And you could really just cater your practice to what you wanted. You could do everything. You could do a small specialty." P6 BC<br>"I felt that that push and that obligation from the medical school that it's an important thing to work rurally, it's an important thing to be a comprehensive generalist, like it was a really huge mandate in the medical school. And like I still definitely feel that now. Like if I did give up my family practice to do something more focused, I would definitely have guilt about kind of like letting down like my mentors, you know." P39 NS<br>"In my experiences with family medicine, in particular, however, I'm not sure if I loved it because it was so fee-for-service volume-based. It felt most of the doctors that I followed, you know, would see 30 to 40 patients a day ... And it was exhausting. And I don't think I saw myself in a model like that. And so even though I chose family, I think in my mind I knew I wasn't going to practice in that manner." P61 ON |
| Residency | "Definitely [my preceptor] had like a full scope practice. You know, all ages and everything. And so I liked that aspect of it. But at the same time, didn't want to be on call 24 hours a day and that sort of stuff. So I think that it definitely influenced how I wanted to divide up my time. I mean I'm sure it influences it." P22 NS<br>"When going through residency in [major city], you're kind of a lower totem pole learner... There's a lot of learners. So again, in my residency, my best times were times I was outside the city. So my [small town] rotation for 3 months again just kind of reaffirmed for me that comprehensive family medicine was what I liked. The kind of full care, hospital, office, different setting family medicine was what I really was passionate about." P26 NS<br>"I mostly decided on doing hospitalist work just based on the different rotations I did during my residency." P60 ON |

| Professional influences on choice of practice type or model |  |
| --- | --- |
| Early career experiences | "And I took a locum, a 2½ day locum position at the site where I had done my primary family medicine site during residency. So I just continued on there ... And so to me it was an opportunity ... Knowing that there was going to be ... the transition was going to be stressful and that as a sort of next step, I wanted to do it in a familiar environment with familiar people with whom I could feel comfortable asking questions." P11 BC<br>"I did like a few clinics as well as the hospitalist work. And then I decided I liked just doing hospitalist work." P60 ON |
| Acquisition and maintenance of skills | "And so I started doing the rural locums. So yeah, part is financial, part is just kind of variety, and another part is also just making sure that I'm still competent, that I'm able to like keep up my family practice licence." P18 BC<br>"And so it's like every skill set, you know that if you don't keep using these things, they're going to be gone. You're going to not have those skills in a while. You know, like it's like that expression – use it or lose it ... I very much thought I've got to keep this stuff alive. So I've tried with my locums to keep that stuff alive." P35 NS |
| Policies and regulations | "I think the fee-for-service system in this province is broken. And the only way that you could keep up with other professional options is to do kind of crappy medicine. And it's disheartening to try to choose between making a more reasonable income versus doing a good job. So I didn't make that choice. I picked something else [focused practice]." P5 BC<br>"I was going to take a job at a community health centre ... doing a blend of addiction medicine and primary care. Which I would have been very happy doing. And then the provincial government took away the funding. So that's not happening anymore." P45 ON<br>"And the ambiguity as to how contract negotiations are going and that relationship has dissuaded me from considering signing onto a practice until I really know what the lay of the land is going to be like." P48 ON |
| Practice management responsibilities | "... family medicine clinic is more exhausting than it sounds like it should be. There's a huge burden of paperwork. And there's a huge burden of demand with care ... And so just for me, I can't do family medicine clinic 5 days a week." P26 NS<br>"I actually don't see the financial benefit in opening up my own practice unless I'm able to employ plenty of doctors. And given all of the job postings online for family doctors to work at practices, and the really low overheads that people are offering because it's so hard to find a family doctor to work with them ... It is not a risk I'm willing to take right now." P59 ON<br>"You know, if you start your own practice, and then you want to go on holidays, and you can't find a locum because no one wants to work for less money than they would get somewhere else, then you're kind of in a bit of a bind because you can't leave." P62 BC |
| Desire for control | "I guess right now just that sense of not wanting too much commitment and wanting autonomy over my schedule after seeing many colleagues that feel chained to work ... I think the past 5 years influenced my inability to commit to a job and stay mostly as a locum, and try to keep lots of freedom in my schedule." P6 BC |
|  | "And so from that perspective, you have more control in a situation like mine [solo practice] where you're able to fine-tune things exactly the way you want them to be. And something like that might not seem very big. But if it's something that bugs you every day that you work, you know, for the next 25 years, it could rob you of the joy and satisfaction that you derive from work." P46 ON |
| Meaningful and valued work | "So in the [low-income neighbourhood], for example, I like that I'm not doing fee-for-service there because it gives me the ability to spend more time with my patients. I think that they need it. For example, it's not uncommon to have somebody coming in in crisis and suicidal, or something like that, or having some sort of a family crisis. And in a fee-for-service clinic, it's impossible to take an appropriate amount of time with a patient like that." P2 BC<br>"... there are aspects of office-based family medicine that are rewarding but it's not always very interesting. So the relationships you build with your patients are rewarding. But the actual day-to-day work of office-based family medicine I find becomes not that interesting or engaging over time. So I'm happy to do it as a part of my practice mix but realize that doing it as a 40 hours a week, I become bored with it." P37 NS<br>"I love the opportunity that academic medicine and education provides ... I get to do all kinds of things. I do clinical medicine, I do didactic teaching, I do some leadership, administrative stuff. I can do research. So I think career-wise and for my own mental health, being able to have your hand in a bunch of different pies is really interesting." P63 ON |
| **Personal influences on choice of practice type or model** |  |
| Financial considerations | "Like really we have to do what pays well because otherwise the debt seems insurmountable. So I was seeking out things that paid well. Like my northern locums and the hospitalist pays better than family med clinic. To be honest, everything pays better than clinic. So I am not surprised people don't choose it initially often. And so that's a big consideration for me." P26 NS<br>"Medicine is ridiculously expensive. I'm not someone from a particularly rich background. And so it certainly has been a significant debt for medical school. And I'm trying to claw my way out of it as fast as possible. Which is challenging. I think it might be part of the motivation to continue to do like the rural locum work." P52 ON |
| Family and relationships | "I don't do any emergency medicine but it's something that a lot of family doctors do here, and would have been an option. Or doing some extra walk-in clinics in the evenings, I've definitely forgone those to be at home and help support my wife and just to be home with my little one in the evening and get her tucked into bed." P23 NS<br>"Because I don't have a partner, it's given me the flexibility to explore doing locum work and then signing on in the community." P55 ON |
| Gender | "I think patients have different expectations of male physicians and female physicians. And that plays out in different ways, I think. But in terms of we were talking about compensation and so on, I think in a fee-for-service type of environment, that's not beneficial to female physicians ..." P2 BC |
|  | "The prospect of having to find a maternity leave locum with the difficulties that that entails ... is a little bit frightening, I'll put it that way. And it discourages me thinking about joining a model like that [community-based practice] when I'm worried about being able to find coverage or interrupt the continuity that I would want to provide to my patients." P48 ON |
| Personality and personal interests | "I like variety and I like diversity and I'm a bit of a ... I'm a traveller and like to be on the go. So I think that's a big part of why I'm working different places and coming and going from these various clinics and work settings." P12 BC<br>"I'm indecisive and I'm curious. And so that's one of the reasons why I continue to locum and I've enjoyed locuming. Well, the indecision part is that I don't really feel ready to settle into a practice of my own just yet because I really want to get a sense of who I am ..." P48 ON |
| Work-life balance | "I think that family medicine, it can be a pretty thankless job these days. And the main thing that is attractive to me about locuming is the ability to work when I want to and take time off when I want to. And my colleagues I see taking on practices, particularly early in their careers, are burning out pretty quickly." P2 BC<br>"Why would I go and work as a family doctor, see patients for 6 or 7 hours, have 3 hours of unpaid paperwork on either end of it, and make, you know, \$600, \$700 a day when I could go work as a hospitalist and make \$1,500 a day? I can go work as an Emerg doctor and make \$1,500 a day. I could go do my work, go home, be with my family, and not have to worry about any of that. ... I've made my career to fit my life, and not the other way around." P14 BC |

## 1. Practice Type and Model

In our analysis, ‘practice *type’* refers to community-based practice, focused practice, or locuming; ‘practice *model’* refers to solo practice, group practice, or region-specific payment or practice model (e.g., Family Health Teams in Ontario). Illustrative quotes are presented in Table 5.

### Training influences on practice type and model

Participants who reported enjoying their **medical school** rotations or seeing mentors demonstrate the breadth and depth of family practice said this influenced them to have a community-based practice of their own. Some noted that their medical school had an explicit focus on community-based practice and that this instilled a sense of responsibility to work in a community-based practice. Others learned from medical school mentors with a niche practice, and reflected this by offering focused services themselves (e.g., emergency, sports medicine, mental health services). Exposure to the challenges of the FFS payment model during medical school sparked a desire for alternative payment models (e.g., salary or capitation).

Some mentors and influential people in **residency** highlighted the benefits of community-based practice over working in more focused settings (e.g., emergency) and generally encouraged participants to explore a variety of settings. Numerous participants reported emulating their preceptor’s practice after residency. However, in some cases, preceptors’ large work volume actually discouraged participants from pursuing community-based practice. Participants perceived that urban placements offered fewer opportunities for hands-on training due to the availability of specialists, and this reinforced their interest in rural community-based practice. Those who had rural residency experience reported that they continued to take on rural locums after residency to maintain the skills needed for community-based practice. Rural residencies were also reported to include hospital-based practice (e.g., hospitalist care, inpatient care, obstetrical call), which some enjoyed; others found it stressful and chose to abandon that part of practice after training.

#### Professional influences on practice type and model

**Early career experiences** after residency, when participants tried different practices, were reported as strongly influential. Some participants used locuming to find a practice they wanted to join long-term; others built a roster of locum sites through which they enjoyed rotating. Participants reported that wanting to **acquire and maintain their skills** influenced their decision to locum, to have a community-based practice, or to complement this with work in emergency and obstetrics.

By contrast, **policies and regulations** influenced people away from community-based practice. Specifically, FFS, low compensation, and restrictions on joining certain practice or payment models were all reported as key factors in the decision to start a focused practice. Participants felt the FFS model was not conducive to providing quality services (e.g., it restricted appointment length times) and caused income instability/unpredictability. This was compounded by the perception of increasing costs relative to fixed fee codes and a resulting dissatisfaction with the level of compensation. The perceived amount of unpaid work in community-based practice further reduced physicians’ desire to open a new practice. Some participants sought alternative payment models that they felt facilitated patient connections and continuity; however, limited opportunities for positions in certain practice models (e.g., Community Health Centres in Ontario), restrictions on entering capitation-based models, and perceived constraints in contract positions caused some participants to turn to locuming or focused practice instead. Several participants were concerned about the future of family medicine (in particular due to the uncertain relationship between family physicians and provincial governments and changing health care policies) and said that this influenced their decision to avoid community-based practice.

The added workload associated with **practice management** – including administrative tasks, billing, and human resources – was reported by participants as a disincentive to work in a community-based practice. In particular, participants reported difficulties finding locums to cover their practice when needed, identifying fellow physicians to join their practice, and having inadequate business training.

A **desire to have more control** over practice management, schedule, and practice style influenced decisions on what type of practice to pursue. Others focused on **meaningful and valued work** such as providing diverse and intellectually stimulating services in acute care settings or as locums. Participants expressed a desire to spend more time with patients during appointments, have the time to build therapeutic relationships, and ensure patients have continuous access to services. These preferences caused some participants to select capitation-based practices or collaborative practices when they were available.

#### Personal influences on practice type and model

Some participants remarked that **financial considerations** such as uncertainty in their personal financial situation, debt, or need for additional income made them reluctant to start their own practice; instead, they pursued the most lucrative options (e.g., locums, focused practice). Participants shared the role that **family and relationships** played; being single and/or not having children gave them opportunities to take on various types of practices or locuming, while those with children reported feeling more limited in their choices. Having children reduced interest in emergency medicine, inpatient medicine, and evening walk-in practice, while increasing desire for salaried models that reduced take-home work. For others, difficulties finding locums for planned maternity leave and perceived judgement by patients that maternity leave was a **gender**-related influence on decisions to focus practice. Personal discomfort with patients’ gender-based expectations about physician behaviour, including expectations that women physicians would provide more emotional support than men, discouraged some from community-based practice.

**Personality and personal interests** were likewise reported as influential. Those who enjoyed travel, an active lifestyle, and were comfortable with change and uncertainty reported interest in locums or work across many settings. By contrast those participants who were not comfortable with uncertainty were drawn to a focused practice.

Participants highlighted that **work-life balance** was an important consideration in their choice of practice model; however, there was no consensus on how to achieve it. Locuming was perceived as a way to gain schedule flexibility and a manageable work commitment for some, but others feared burnout from locuming and thus chose to start their own practice. A desire for flexibility generally influenced participants away from a community-based practice. Solo practice was perceived as a model associated with high work volume, need for around-the-clock availability, and difficulty finding coverage for time off. This led some participants to instead choose team-based care models. Hospitalist practice was either perceived as allowing for flexibility and better hours, or as requiring overtime and potentially leading to burnout if combined with community-based practice.

#### 2. Scope of Practice

‘Scope of practice’ refers to current areas of clinical practice, populations served, services delivered (in community-based or focused practice), and extra-clinical activities (e.g., research, teaching). Illustrative quotes are presented in Table 6.

**Table 6.** Participant perspectives on influences on scope of practice.

| Influence | Quotes |
| --- | --- |
| **Training influences on scope of practice** |  |
| Residency | "And so I probably got the best person I could have. She was a young female physician, had done emergency medicine as well as family practice. And she also did the minor procedures clinics ... Honestly, my practice is set up to mirror hers almost identically because that's how much I liked it." P41 NS<br>"I did it [residency] at the [clinic name] family health team. It was an academic unit. We saw a lot of complicated patients. I felt very prepared by the end of it to start working in full spectrum family practice." P49 ON<br>"I don't feel comfortable doing obstetrics, for example, because my exposure wasn't quite the level that it would have needed to be for me to be able to walk out of residency and feel ready to take on delivering babies all the time. So that certainly shaped how I practice." P52 ON<br>"I felt more comfortable even moving back to more of an urban setting and doing emergency medicine and obstetrics. The fact that I did have the rural practice skills really gave me the confidence to still continue to pursue it, even though it's uncommon for urban family physicians to continue to do obstetrics and emerg." P58 ON |
| Additional training | "And then I did a third year fellowship. And so at that third year, I did more focused training in women's health. So I did some work with women living with HIV. I did work around menopause, and sort of different life cycle issues in women's health. I became trained to provide abortions and do focused sort of sexual and reproductive health work in [current practice]." P1 BC<br>"I ended up doing some extra training in obstetrics which like opened up some doors for locums in my community for when I finished. ... A lot of it I think is to support rural physicians to do extra training so they can go back to their community and be more comfortable. I was able to do it ... to become more comfortable practicing OB." P7 BC<br>"That's [sports medicine] a fellowship trained role. There's different ways in. They try to close the door on you so you can't get in. So I'm not even allowed to do it and write an exam for two years after I get out." P50 ON<br>"It seems like right now in order to do full-time palliative care in many areas, you need to have a PGY3 in palliative ... I'm told my skills are not adequate to run a palliative care unit or to see palliative care patients. I find that frustrating. The idea of doing a PGY3 feels a bit daunting." P54 BC<br>"I was actually planning on doing my third year fellowship in obstetrics. And I ended up not doing it because I had my daughter and I didn't want to be away from home. But instead of that I chose to do a locum with family medicine obstetrics support. So then I kind of, you know, used that as to further develop my skills in obstetrics during my locum time." P58 ON |

| Professional influences on scope of practice |  |
| --- | --- |
| Early career experiences | "I had a couple of careers before I came in. But primarily working in community health for 20 years... And a master's degree in social work before I came to medicine like kind of by fluke. Like all of that influenced where I'm at. ... And I knew that I wanted to work with marginalized communities and complex patients. And that's effectively what I'm doing. So I'm very interested in helping folks who are experiencing a lot of barriers to healthcare, and understanding how, for example, the social determinants of health impact patients' ability to be healthy and have contented lives." P11 BC<br>"When I was in high school and I guess in undergrad as well, I did a lot of work at a nursing home. I did volunteer work at a nursing home. And I felt that that really actually had probably influenced my decisions to work as a hospitalist in family medicine, because I was always so exposed to the age group which I'm working with now." P60 ON |
| Meaningful and valued work | "Like I really want to be doing something that feels meaningful to me, and feels that it's in line with my values and ... Like it sounds super cliché or whatever but I want to be able to feel like I'm making some kind of difference, and that what I'm doing is important." P29 NS<br>"I think I like to kind of build some close relationships with my patients. And I think that's part of the reason why I like palliative care – because you kind of get to spend more time, kind of longer appointments with people, and you really kind of get to know them and what they're about and what their life is like." P51 ON<br>"Our obligation is to be responsive to our communities ... So I have my favourite things in medicine, and then things that really I end up having to hone just by the population in the community that I'm serving." P56 ON<br>"And working I think more with a marginalized population. A population where you felt that you left and you said, okay, if I wasn't here, these people might ... I don't want to say not gotten care because there's definitely lots of staffing there. But that they might face barriers in different ways. So that's kind of the population I kind of like to work with." P61 ON |
| Confidence with service provision | "I don't do IUD insertions. I know that's really specific. I don't have enough of a patient population that wants them routinely enough to maintain my competency." P41 NS<br>"... I feel very, very comfortable with palliation. We see that a lot at the hospital. And that's not really an interest but I find a lot of physicians are not adequately trained. I know I'm not but I think I'm more trained than some of my colleagues that I work with unfortunately. " P47 ON |

| Personal influences on scope of practice |  |
| --- | --- |
| Family and relationships | "We're fortunate in that my husband's a medical specialist. And so I feel very little burden now in my career ... I feel less burden from a family perspective to bring in income ... in my marriage, obviously I've chosen to work less—we made that as a partnership decision—than had I been a single person or primary breadwinner." P19 BC<br>"It's [obstetrics] certainly like my all-time favourite thing to do in family medicine. But my husband is an obstetrician. And realistically balancing two call schedules with family is just ... it would be a bit of a nightmare. Now once the kids are lot older, perhaps I would get retrained and consider doing that again, considering how much I love it. But realistically I don't know that it's ever going to happen again." P36 NS<br>"I myself have a 3½-year-old. So before, like when I was first in practice for that first year, I probably did more stuff outside of my office. Like doing obstetric call. But whereas overnight, I've done less of that since my son has been around. And I would say likely when he is older, I would probably pick up doing more of that. " P47 NS |
| Gender | "So my preceptor, we used to get procedure clinic every month. And I loved it and I was good at it. ... Then I went on mat leave ... Then I was away for over a year and a half with my second child. And then doing locums, I wasn't doing a lot of procedures because you're sort of bouncing around a lot. ... Like by the time I got to [current practice], I think I'd been 2 years or even 3 years since I'd done a procedure on someone. So I lost like my own confidence." P22 NS<br>"So with gynaecology, for instance, I always give patients the option of, for instance, seeing me for that issue or seeing another physician – a female or whichever gender they choose. That would have affected me in terms of doing gynaecology versus anything else. And I think it does have bearing for me as well. Even though again I do the prenatal assessment, I usually don't follow people prenatally. I do believe it has some bearing there ... just for me it's more I have this belief that a woman would be more comfortable around another woman for that situation rather than a man or any other gender." P33 NS |
| Personality and personal interests | "Like this very traumatic, terrible case happened. And as awful as it was, like I think it showed me that I ... had the skills and the bedside manner to sort of help people in those times of need. ... I practice with my emotions. Which is good and bad. But I really connect with people that way. And I think before it kind of scared me about doing things like palliative care and emergency medicine. But I think it showed me that I could do those and that it was something I wanted to do." P17 BC<br>"I'm extremely patient. And in a lot of family medicine, you do get a lot of geriatrics. But I do feel like I have a little bit of a leaning towards geriatrics. And the reason I do feel you need more patience than any other is because just in terms of how information is communicated now or how the geriatric population does need more time. That's kind of why I do sort of do long term care as well." P33 NS |

### Training influences on scope of practice

**Residency** experiences were reported as being associated with either an increased scope of practice – when preceptors provided a diverse range of services and increased participants’ competencies in these services – or a decreased scope of practice when training was lacking. **Additional training** enabled participants to provide certain services (e.g., emergency medicine) or care for populations with specific needs (e.g., people with substance use disorders). Some participants described not being able to obtain additional training, which limited the services they could provide in their practice (e.g., obstetrics, palliative care). However, some found workarounds to gaps in training, such as working with other physicians to gain experience or learning on the job.

### Professional influences on scope of practice

**Early career experiences** were reflected in some participants’ practice choices. For example, previous non-medical degrees or career experiences increased participants’ interests in mental health services, social determinants of health, or specific patient populations (e.g., equity-deserving groups). Some participants reported that their early career experiences in rural settings increased their desire to offer a broader range of services.

Participants reported that their scope of practice was influenced by a desire to engage in **meaningful and valued work**. Their desire to have a positive impact on their patients’ lives influenced their decision to offer specific services such as sexual health care, palliative care, or obstetrical care. Similarly, they valued meeting patient and population needs, which influenced participants’ scope of practice as they worked to fill service gaps in their community (e.g., medical assistance in dying, opioid agonist treatment) or support particular patient populations (e.g., women facing barriers to care, transgendered people). Teaching was incorporated by some to ‘give back’ to the next generation of physicians.

Reduced **confidence** in the delivery of specific services or care for specific populations led some to limit their scope of practice. Confidence was influenced by time away from providing certain services (e.g., minor surgical procedures) or a lack of volume (e.g., IUD insertions, obstetrics). When participants felt confident with specific populations or services, they were more inclined to include those in their practice.

### Personal influences on scope of practice

**Family and relationships** were reported as very influential. Participants with children chose to not offer certain types of services, such as hospitalist or obstetrics care, due to unpredictable schedules or night shifts. For some, having a spouse with more lucrative employment gave them the freedom to pursue a scope of practice that was in line with their interests rather than with higher income. For others, however, having a working spouse meant they had more family responsibilities, which restricted their ability to provide full-time community-based care or obstetrics services.

Participants also reported that their **gender** was sometimes influential, particularly when faced with gender-based expectations about physician behaviour. The perception that patients prefer women physicians for women’s health issues influenced some men physicians to not provide those services.

Participants reported that taking maternity leave reduced their confidence to provide certain services due to time away from active practice. Lastly, participants spoke about their **personality and personal interests**; some said that their empathy and patience led them to work with certain populations (e.g., geriatric populations).

#### 3. Location of Practice

We define ‘location’ as the geographic location of participants’ practice or as the distinction between rural or urban practice. Illustrative quotes are presented in Table 7.

**Table 7.** Participant perspectives on influences on location of practice.

| Influence | Quotes |
| --- | --- |
| **Training influences on location of practice** |  |
| Medical school and residency | "My family medicine rotation in the city compared to what I did here [rurally], like just the collegiality between the physician and the patients was so different, I thought. Anyway, I really enjoyed it. And I knew I was going to do rural family medicine just from that point on." P28 NS<br>"I did a lot of my med school family rotations rurally around [names of cities] and smaller towns around that area. And I think they all just reaffirmed for me that it was the right setting for my practice in the future." P44 ON |
| **Professional influences on location of practice** |  |
| Policies and regulations | "So I've noticed, you know, if I work as a locum in a place and I don't feel like I'm being compensated well, I don't go back to that place. And part of why I've been working up north is also because it's compensated a bit better than [city] was." P12 BC<br>"So like overall, working in BC has been very stressful. Like the amount of work for the compensation is a lot higher than what I'm used to practicing in Alberta. And just going back to BC where BC is notorious for having like a culture where you have to pump through patients whether you want to or not, that work is very tiring. And to kind of give myself a break where I can slow down, I go to Alberta." P21 BC<br>"To me the licencing process was incredibly cumbersome going from one province to the other, and it's definitely a barrier. I've had job offers in other provinces. But the idea that I'd apply for another licence definitely throws a barrier to why I don't want to work in any other provinces." P54 BC<br>"So when I looked at other compensation models both in Ontario and outside of Ontario, in different provinces and territories, I realized that Ontario is a very good place to work. Specifically rural Ontario is a very good place to work." P55 ON |
| Challenges in rural settings | "I think it's more in terms of being kind of worried or afraid of not being able to provide the care that is needed [in a rural setting] because you don't have all the supports that you have in an urban setting. So here if someone kind of breaks something, you just call the orthopedic surgeon or send them to the ER and they'll deal with that. If you are in a very small community, you probably have the support over the phone. But at the end of the day, you're still the one who's doing all those things that I don't feel comfortable doing." P4 BC<br>"However, when we talk about more rural settings, because I was trained in a more urban program, sometimes I feel like the skills to manage acute situations in a rural setting, the program may not have given me." P16 BC<br>"And it was kind of a little bit heavy in terms of workload [small town in Newfoundland] and trying to balance that with life. So we looked at other opportunities. Which is what brought us to NS." P39 NS |
| Meaningful and valued work | "I like having a versatile skill set in medicine. Like I don't know that I would be very happy with a very narrow scope of practice. ... And a rural family practice allows for that especially because there's a lot more opportunities to kind of get skills outside of the usual." P39 NS<br>"I chose to do rural practice because I wanted to have family medicine skills but also other skills that could make me a good doctor overall." P58 ON |
| **Personal influences on location of practice** |  |
| Family and relationships | "Yeah, maybe if I'd married someone different, I might have ended up doing rural work. But yeah, I ended up with a fairly urban partner. I think if it was earlier on in my career, I would have considered doing rural-type work. But I don't think that's in my future." P9 BC<br>"I think the work that my wife does and where she works plays a large role in me ending up geographically where I am. And I think that also plays a role in where we're living. So I think that's really influenced where I practice and my style of practice." P13 BC<br>"... moving back home allowed us to be close to both sets of parents as well as siblings and their children. As well as live and work in a city that we know very well but also is much more affordable and allows for a more balanced way of life." P35 NS<br>"So partner-wise, you really can't go anywhere rural because your partner has a job in a city. And that kind of limits where you can move or work as well. Because you can't go to a rural area if they do something that's more like big business or like a big kind of executive thing. You can't go to a small town." P50 ON |
| Community | "The BC College has very strict guidelines around what kinds of personal relationships you can have with patients ... and in the sense that, you know, you can't be friends with your patients, you can really have very minimal sort of personal interaction with patients. Which in a community of a 1000-2000 people where you're the physician to essentially everybody in the community means that it's not a place you can really live a life. So I think the only way to work in a place like that is really to come and go, and have a life elsewhere." P2 BC<br>"And the longer that we've stayed in [name city] ... the less we are likely to leave because we've made these really great friends both personally and professionally ... So I guess having gone to school and done residency there has definitely shaped where we've decided to stay. Because now my husband has worked there and made friends ... and [we] have this whole community around us." P6 BC<br>"I trained here, I know the system, I know all the people, I know how everything works. I know who to go to when I have a question ... why would I want to go jump straight into an unknown situation when I already know how everything works? So it just was like I already know how this all works so why not just start working?" P43 NS |
|  | "I kind of had a plan after third year of medical school ... And I knew where I was going to be, at least close to where I grew up. And there was good opportunity about 45, 50 minutes from the town I grew up in. So I kind of just went there automatically." P44 ON<br>"But it's a really wonderful feeling to be going through the community, whether you're walking on the street, in a restaurant, a grocery store, and people come up to you and say hi, and they ask you how you're doing. And they don't ask because they have to. It's because they care. Because it's that sort of community." P55 ON |
| Personality and personal interests | "My lifestyle is vastly based in the outside. And I couldn't get outside as much as I want in a city. And I just figured out what things truly make me happy and the things that I truly enjoy, and then worked to be in a setting that allows me to do them." P44 ON<br>"I loved the lifestyle. I loved how beautiful this place was. And it was those factors that made me decide to apply for my full licence here." P54 BC |

### Training influences on location of practice

Participants reported that having **medical school** electives or **residency** placements in rural, small town, and northern practices resulted in a greater desire to work in those locations, in particular when mentors modelled community-based rural practice or encouraged the participant to join their rural practice after graduation. Participants noted that their residency placements either allowed them to get familiar with a location and want to stay, or influenced the choice to not work in the area.

### Professional influences on location of practice

Participants reported that their choice of practice location was influenced by **policies and regulations** that impacted work volume and pay. For example, opportunities to join a payment model that aligned with their preferred style of practice, provided higher income, or required no overhead payments influenced the decision to work in certain locations (e.g., rural locums). Provincial licensing regulations were reported as costly and deterred participants from working in more than one province. Those who did get a second provincial license described low remuneration in their home province as a factor in their decision.

Some participants shared that they moved to an urban setting because of the **challenges they faced working in low-resource, rural communities** due to high work demands. Similarly, others felt uncomfortable working in rural settings because they lacked confidence to offer a broad scope of services or to be the only physician in a region. Those who were uncomfortable working in emergency departments noted it as a limitation to their ability to locum in smaller communities. Others reported that rural locations provided an opportunity for **meaningful and valued work,** including gaining and maintaining a broad range of skills, pursuing intellectually stimulating work, having variety in practice, and finding meaning working in under-resourced settings.

### Personal influences on location of practice

Participants shared that they worked in or near their childhood community because of **family and relationships**. This was emphasized by participants with children, who described wanting to be near family and in a community with a strong education system. The desire to live close to aging parents and concerns about future caregiving responsibilities were other considerations in choosing a practice location. Many participants also noted that their spouse influenced their decision about where to work, such as when a partner did not want to live in a rural area. In other instances, participants worked where their spouse had their residency, practice, or other work site. Participants reported that rural **communities** reduced opportunities for friendships and relationships. However, others found that being familiar with a community and feeling appreciated influenced their decision to stay there. Participants also reported being drawn to communities that aligned with their **personality and personal interests** (e.g., outdoor recreation).

#### 4. Practice Schedule and Work Volume

We define ‘practice schedule’ as both timetable and time allotted to practice. ‘Work volume’ is defined as the workload associated with participants’ practice. Illustrative quotes are presented in Table 8.

**Table 8.** Participant perspectives on influences on practice schedule and work volume.

| Influence | Quotes |
| --- | --- |
| **Professional influences on practice schedule and work volume** |  |
| Offer better access to care | "And how we set it up is because there's enough of us to do this, one person each week will take after 10:30 in the morning so that all of their appointments would be open. So they'd be available to any patient that's part of our practice for a same-day visit. So it's not walk-in style. If patients call and need to be seen urgently, like for something that was infectious or if there was a last minute refill or that sort of thing then we take turns each day to see them." P42 NS<br>"I have a lot of patients who work. So I just decided to kind of change my hours to accommodate a lot of my patients who prefer to see me in the evening." P51 ON |
| Remuneration | "Like ideally for me I would do like ... 75% hospitalist work, 25% in an office, doing family medicine in an office. Hospitalist work I find a little more interesting. It's convenient that it also pays quite a bit better. If it didn't pay as well, would I ... Like there's a combination of things. Why do you pick up work? Part of it's how much you enjoy the work, part of it's how well compensated you are for the work. ... You can't always parse it out precisely as to how much each of those factor in. But they both play a role." P37 NS<br>"If I have 30 people scheduled in a day, and say 10 people decide not to show up, that's 10 patients who could have been in my office. That's also a significant loss of income [in a FFS model]. So if I'm not guaranteed patients to show up on evenings or weekends, there's no incentive for me to open my office because I will actually lose money by doing that." P41 NS |
| Acquisition and maintenance of skills | "Because it's very hard in a newer practice for a physician ... Everything is new and everything is challenging. ... And just my baseline stress level of seeing those patients is going to be a ton higher because I'm just less confident about everything. And so it's just really hard to work at full-time levels with that underlying increased stress. ... I think the idea of introducing yourself into that more slowly with lots of time off is a very important thing." P15 BC<br>"Yeah, I mean I don't think it's sustainable forever [working a 6-day week]. You know, I think right now as like a brand new grad, I want to solidify my skills. And I'm still trying to figure out exactly sort of what my ideal practice looks like. So try and keep my skills up and trying to do all these different things so I don't limit myself in what I sort of feel comfortable to do." P17 BC |
| **Personal influences on schedule and practice volume** |  |
| Work-life balance | "[I] have a lot of interests outside of work. So I want to be able to have the time off to engage in those interests. Like for example, you know, I snowboard a lot. And so I try not to take too many rural locums like during the snowboarding season. But then ... when I'm not doing much snowboarding or hiking then I work lots. So that allows me to make a bit more money." P18 BC<br>"So that was a positive thing – to realize that I didn't have to work 5 days a week, you know, 50 weeks a year to earn a living. But that I could work part-time and still maintain a lifestyle that allows me to focus my leisure activities or personal activities a bit more than just, you know, grinding away at a job just to pay the bills." P24 NS |
|  | "The reason I've had to cut back on hours is I was finding I ... would put my kids to bed and ... I'd eat supper and then spend the whole rest of my evening doing paperwork. So it just wasn't feasible to continue doing that for the long term." P36 NS<br>"I kind of got tired of the hospital inpatient setting. I think maybe in the future, like much later down in my career, I might want to come back to that. But right now I don't really want to be on call and spend long hours in the hospital – overnight hours, things like that. So I did want more of a bit of 9 to 5 structure." P38 NS |
| Financial considerations and pressures | "Like I have worked 6 days for several months. Really bad weeks are 7 days. But some weeks can be down to like 3 days a week. So it varies. ... Now that I'm approaching the next stage of my life, I hope to be less busy. But with some of the financial pressures with debt and housing, especially in BC, sometimes you just need to work more ... in BC ... I think the realization that, you know, even though physicians are well paid, you know, it still can't buy you a house or even like a condo that's big enough for a family." P21 BC<br>"So I am the primary breadwinner in my house. I'm married. My husband would make like just kind of like an average income, I guess. So I'm basically ... There's a decent amount of pressure on me to work as well." P34 NS |
| Family and relationships | "I have two kids, another one on the way. And I've tried to make sure that I don't work too much. That I have that balance and that I have enough time at home ... But I think if I didn't have a family I probably would work a lot harder and do different things, and getting more involved." P51 ON<br>"I guess now definitely having a new ... like a young child, that's definitely changed things a lot. Like I'm much more restricted in hours that I can work or that I want to work. I want to work less." P62 BC |

### Professional influences on practice schedule and work volume

Participants shared that wanting to **offer patients better access to care** influenced their clinic hours and work schedule (e.g., open in evenings, working more). **Remuneration** considerations also influenced how participants allocated work time. For example, the ability to earn a higher income as a hospitalist influenced some participants to consider reducing their hours in community-based practice and spending more time working in the hospital. Working in a FFS model was cited by some as a reason to not work evenings and weekends, since there were more no-shows for which they would not be paid. Some participants also shared that wanting to **acquire and maintain skills** either influenced them to work more days per week (i.e., to practice the skills) or to reduce their work volume until they felt more comfortable providing a specific service.

### Personal influences on practice schedule and work volume

Participants noted that **work-life balance** considerations influenced their schedule and work volume: some adjusted their schedules around personal interests or family while others reduced work volume, built in rest time, opted out of night shifts, or kept their schedules flexible to avoid burnout.

**Financial considerations** led some participants to increase their work volume. **Family considerations**, such as parenting responsibilities and limited childcare availability influenced some participants to decrease work hours and/or prevented them from working weekends and night shifts.

## Discussion

This qualitative study of 63 early-career family physicians identified how training, professional, and personal influences were reported to influence choice of practice type or model, scope, location, and practice schedule and work volume. Multiple factors were reported by each participant, resulting in a complex set of often interacting factors that influenced individual practice choices. Training was reported as influential for all these practice characteristics, except schedule and volume. Participants described the influence of rural training experiences and the important role played by mentors and preceptors in medical school and residency. Professional factors influenced all practice characteristics; policies and regulations and a desire for professional satisfaction were commonly mentioned. Personal factors, in particular family responsibilities, work-life balance, and personality or lifestyle were reported to influence all practice characteristics.

While many studies have examined factors that influence medical students’ choice of family medicine ^22, 23^, few have examined choice of practice characteristics among active family physicians. Those that have often focus on a single practice characteristic, such as scope ^3, 24–26^ or practice type and model ^27^, and few examine early-career family physicians ^24, 27^. These studies confirm our findings of the influential nature of training and mentorship ^3, 25–27^, early-career experiences ^25, 26^, work-life balance ^3, 26, 27^, professional satisfaction ^3, 24–27^, family ^3, 26^, policies and regulations ^3, 26^, and remuneration ^24–27^.

Our findings, however, did not always align directly with those in the literature. Likely due to differences between health care systems, Russell et al. identified more factors affecting practice choices among American physicians at the institutional level (e.g., choices made by organizational leadership about service provision) ^3^, while we identified more factors related to provincial policy. A study by Gosden et al. in England found that new family physicians were sometimes averse to practices in areas of high deprivation ^28^. Some of our participants instead highlighted the professional satisfaction they gained from meeting community needs and working with equity-deserving populations.

Family physicians face tensions between caring for a patient population with increasing complexity and the growing administrative burdens and costs of community-based practice ^29, 30^. For some, including some focused practice services helped create a more sustainable career ^31^. Given the complexity of often interacting factors in our study’s findings, governments that want to address workforce concerns and increase the availability of comprehensive primary care must look at modifying multiple factors that create barriers to community-based practice. This includes policies and regulations, but also factors such as physicians’ desire for meaningful and valued work, work-life balance, and meeting the needs of patients. Lastly, factors such as family responsibilities, friendships and relationships, and personal interests should not be ignored.

A strength of this study is its breadth and open-ended nature. The use of qualitative enquiry allowed participants to describe the most relevant factors that shaped how they set up and maintained their practices. Given the broad nature of our research question, our interviews did not allow for an in-depth examination into any particular practice characteristic or influence. While study participants worked across different provincial systems, its Canadian context may mean the results do not directly apply to other health care systems.

## Conclusion

These rich qualitative data demonstrate the complexity of factors which shape the way early-career physicians practice. Policies should take this complexity into account; overly simplified policies that target a single factor such as training are unlikely to reverse the trend away from comprehensive community-based practice. Meaningful system reform will be better supported with a nuanced and comprehensive understanding of the variety of influences informing how family physicians choose to practice.

## Data Availability

The data sets generated and/or analysed during the current study are not publicly available due to participant confidentiality and research ethics board requirements.

## Acknowledgements

We would like to thank all of our participants for their valuable contributions to this study.

## Ethics Approval and Consent to Participate

This study received ethical approval from the Simon Fraser University Office of Research Ethics with harmonized approval from the University of British Columbia, the University of Ottawa, the University of Western Ontario, the University of Ontario Institute of Technology, and the Nova Scotia Health Authority. All participants provided consent to participate.

## Availability of Data and Materials

The datasets generated and/or analysed during the current study are not publicly available due to participant confidentiality and research ethics board requirements.

## Authors’ Contributions

AG, MRL, DR, LJG, and EGM conceived the project and led grant submission. All authors contributed to the development of recruitment strategies and data collection tools. AG, LJG, and EGM supervised data collection. ER, LJ, and MMcK conducted the interviews and coded the data. AG, ER, LJ, and LJG analyzed the data. AG and ER interpreted the results and drafted the manuscript. All authors revised the manuscript and approved the final version.

## Abbreviations

● ON: Ontario
● BC: British Columbia
● NS: Nova Scotia

## Appendix: Interview Guide

### Current & Future Practice Description

1. How did your career unfold out of residency?
2. Tell me briefly about your current practice.

- How is your practice o<u>rganized</u>?
- <u>Who else do you work with</u>?
- How are you (and your team) <u>compensated</u>?
- What does your <u>work week</u> look like?
3. Tell me about any particular clinical interests that you have as a family physician.

- Which <u>patient populations</u> are you interested in?
- How have you <u>incorporated these interests</u> into your practice?
- Do you see this <u>changing</u> over time?
4. In what ways, if any, does your <u>current practice differ from your ideal</u> type of practice?
5. How do you see your practice <u>changing over time</u>?

- If IMG with a return of service: how will your practice change after your return of service is satisfied?
6. [If participant mentions “<u>comprehensive</u>” probe: what does comprehensive mean for you?]

### Priorities

7. When you think about your <u>career, what is most important</u> to you?
8. n what ways, if any, did your <u>personal priorities or goals</u> influence your career?

- How did your <u>personal relationships</u> influence your career?
- How did <u>parenthood or caregiving</u> influence your career?
- How did <u>financial considerations</u> influence your career?
- How did <u>your gender</u> influence your career?
- How did your <u>other personal characteristics</u> influence your career plans?
9. (if no exogenous factors emerge in Q 4, 5, 6) What kinds of other influences have you experienced or anticipate that may influence your practice changes over time?

i. E.g. Community, professional, regulatory influences

### Past Experiences

10. How did your <u>medical school experience</u> influence your career plans as a family physician? (*Re-direct away from responses about why family medicine was chosen as a specialty)*

- Positive or negative experiences?
- Did you have any <u>experiences</u> stand out in primary care during training?
- Did any <u>key people</u> influence your plans?
11. How did your <u>residency</u> influence your plans for family practice?

- Positive or negative experiences?
- Did you have any <u>experiences</u> in primary care stand out during training?
- Did any <u>key people</u> influence your plans?
12. Tell me about your C<u>aRMs</u> experience

- Was family medicine your <u>first choice</u>?
- Did you have to make <u>trade-offs between specialty and location of residency</u>?
13. Tell me about any other life experiences you’ve had that influence your career plans as a family physician?

### Why: Policy Environment and Practice Opportunities

14. At the start of the interview, you told me about how you practice now and what you would like your clinical practice to look like. Do <u>you expect you will be able to achieve</u> this ideal type of practice? Why / why not?

- Are <u>opportunities available</u> for your preferred type of practice?
- Are there <u>restrictions or barriers</u> to you having this ideal practice?
- How will you <u>populate</u> your practice?
- How do your <u>gender or other personal characteristics</u> impact your ability to achieve this type of practice?

### Wrap up

15. If you were <u>mentoring a new family medicine resident, what advice would you give</u> them about planning their career in family medicine?
16. Anything else that you think is important for me to know?

